# RT-RC-PCR: a novel and highly scalable next-generation sequencing method for simultaneous detection of SARS-COV-2 and typing variants of concern

**DOI:** 10.1101/2021.03.02.21252704

**Authors:** Christopher Mattocks, Daniel Ward, Deborah Mackay

**Affiliations:** Wessex Genetics, Salisbury NHS Foundation Trust, Salisbury, UK; Faculty of Medicine, University of Southampton, Southampton, UK

## Abstract

We describe a novel assay method: reverse-transcription reverse-complement polymerase chain reaction (RT-RC-PCR), which rationalises reverse transcription and NGS library preparation into a single closed tube reaction. By simplifying the analytical process and cross-contamination risks, RT-RC-PCR presents disruptive scalability and economy while using NGS and LIMS infrastructure widely available across health service, institutional and commercial laboratories.

We present a validation of RT-RC-PCR for the qualitative detection of SARS-CoV-2 RNA by NGS. The limit of detection is comparable to real-time RT-PCR, and no obvious difference in sensitivity was detected between extracted nasopharyngeal swab (NPS) RNA and native saliva samples.

The end point measurement of RT-RC-PCR is NGS of amplified sequences within the SARS-CoV-2 genome; we demonstrated its capacity to detect different variants using amplicons containing delH69-V70 and N501Y, both of which emerged in the UK Variant of Concern B.1.1.7 in 2020.

In summary, RT-RC-PCR has potential to facilitate accurate mass testing at disruptive scale and cost, with concurrent detection of variants of concern.

## Introduction

The Coronavirus Disease 2019 (COVID-19) pandemic has caused detrimental social and economic effects worldwide and, at the time of writing, is responsible for over 2.5 million deaths globally.

The etiopathogenic agent responsible for COVID-19 is the novel SARS-CoV-2 coronavirus. The gold standard test for molecular diagnosis of current SARS-CoV2 infection requires RNA extraction from the given sample followed by real-time qualitative detection of viral RNA by real-time reverse-transcription PCR (rRT-PCR). Whilst different sample types are known to yield different positivity rates through the course of an infection [1], practicalities of sample collection and subsequent processing are the predominant consideration. Generally, RNA has been extracted from nasophyaryngeal swab (NPS) samples collected by healthcare personnel, but more recently saliva has become a popular sample type due to the ease of self collection, though it remains uncertain which sample type is more valid for testing. Attempts have been made to rationalise the test procedure by simplifying the RNA extraction step or eliminating it entirely [2-6]. Such refinements facilitate more streamlined laboratory processing, reduce cost and ease the issue of supply of critical reagents.

Reverse transcription loop mediated isothermal amplification (RT-LAMP) has been developed as an alternative to rRT-PCR [7 8]. Because this reaction uses strand displacement instead of thermal cycling, amplification is extremely fast, and can significantly reduce the time required for the measurement phase of the test. It is worth noting that both rRT-PCR and RT-LAMP rely on indirect fluorometric measurement and as such may require secondary confirmation that the signal has derived from the desired target typically by melt analysis of the amplicon. Expansion of viral testing to community-based or population-based screening is necessary to detect asymptomatic transmission and potentially restore freedom to interpersonal interactions within and between communities.

A laboratory setup suitable for such testing must be capable of handling in the order of tens of thousands of tests per day and the test itself must be sensitive and specific, economical, rapid and robust; and preferably use widely-available and existing infrastructure. Key challenges to scaling include the timely and safe delivery of samples to the testing site, the extraction of RNA, the robust supply of reagents that are in high demand, and the simplification of laboratory testing processes so that they can be effectively automated. To achieve the required scale of testing, a high level of parallelisation is required in the measurement phase of the test.

Typically rRT-PCR and RT-LAMP tests can be analysed in batches of up to 96 samples. Even considering fast turnaround on the analysis platforms, multiple instruments are required to analyse tens of thousands of tests per day.

Next generation sequencing (NGS) represents an alternative platform with the capacity to analyse many tens of thousands of samples in a single instrument run. Individual samples can be labelled with short unique identifying DNA sequences (indexes) prior to sequencing so that the data can be assigned to samples during analysis. The challenge with this approach is streamlining the library preparation process and providing sufficient indexes for the analysis scale required without risk of cross-contamination between samples.

Conventional amplicon-based library preparation methods require separate reactions to amplify the targets and subsequently apply the sample indexes. This process is complex in terms of automation and presents an inherent risk of cross contamination due to the opening and manipulation of tubes containing exponentially amplified target material prior to the application of indexes.

Here we describe a novel assay method: reverse-transcription reverse-complement polymerase chain reaction (RT-RC-PCR). RC-PCR rationalises NGS library preparation into a single closed tube reaction, thus simplifying the analytical process and eliminating risks of cross-contamination. By integrating reverse transcription (RT) into RC-PCR we have developed a single-tube SARS-CoV-2 RNA detection assay using NGS. RT-RC-PCR presents disruptive scalability and economy while using NGS and LIMS infrastructure widely available across health service, institutional and commercial laboratories.

We present a validation of RT-RC-PCR for the qualitative detection of SARS-CoV-2 RNA by NGS in both extracted nasopharyngeal swab (NPS) RNA and native saliva samples.

## Materials and Methods

### Reagents

Synthetic SARS-CoV-2 RNA control was sourced from TWIST Biosciences (MN908947.3). The RT-RC-PCR kit was an accelerated R&D product developed by NEB, incorporating Luna WarmStart reverse transcriptase and Q5 polymerase. The SuperScript™ IV One-Step RT-PCR System from Thermo was also validated for extracted NPS RNA. Oligonucleotides were purchased from Integrated DNA Technologies. NGS reagents were purchased from Illumina and analysis was performed on an Illumina MiSeq instrument using V2 micro flowcells.

### RC-Probe sequences

RC-Probes comprised 5’-[reverse complement of desired target primer]-[universal sequence]-3’. The universal sequence was adapted from illumina sequencing primers published in Illumina document Document # 1000000002694 v01 (February 2016) (https://support.illumina.com/content/dam/illumina-support/documents/documentation/chemistry_documentation/experiment-design/illumina-adapter-sequences-1000000002694-14.pdf). Target primer sequences for the SARS-CoV-2 E amplicon were published by Corman et al [9] and the target primer sequences for the RPP30 human control amplicon were published by CDC https://www.cdc.gov/coronavirus/2019-ncov/lab/).

### Universal indexing primers

Universal primers comprised 5’ -[sequencing adaptor]-[index]-[reverse complement of universal sequence in RC-probe]-3’. The sequencing adaptors were P5 or P7 published by Illumina (Document # 1000000002694 v01 (February 2016)) and index sequences were from the 10bp BFIDT index set [10].

### Samples

Anonymised extracted NPS samples were obtained from Salisbury Foundation NHS Trust Virology department. The standard of care (SoC) diagnostic workflow involved: NPS RNA extraction with the Prepito Viral DNA/RNA 300 Kit (Perkin-Elmer CMG-2017); RT-PCR using the Bosphore Novel coronavirus (2019-nCoV) Detection Kit v2 (Anatolia Geneworks ABCOW5), assaying E and ORF1AB, using the Qiagen Rotorgene as the analysis platform. Extracted NPS samples in which SARS-CoV-2 was detected using SoC testing are here designated ‘positive controls’, and those in which SARS-CoV-2 was not detected are designated ‘negative controls’.

Saliva samples from healthy (nonsymptomatic) anonymised controls were given in the morning before eating or drinking. Aliquots of saliva (100μl) were placed in screw-capped tubes, and heat-treated to 95°C for 5 minutes before storage at −20°C. For the direct saliva test 5μl aliquots were assayed in exactly the same way as extracted RNA. Cultured, UV-inactivated SARS-CoV-2 was a generous gift from Dr Karl Staples, University of Southampton.

### Principle of RT-RC-PCR

Each RT-RC-PCR reaction requires five oligonucleotides; two RC-probes (specific to the amplicon) two universal primers (specific to the sample) and a reverse transcription primer. **Figure 1** shows the logical principle of RT-RC-PCR.

**Figure 1:**
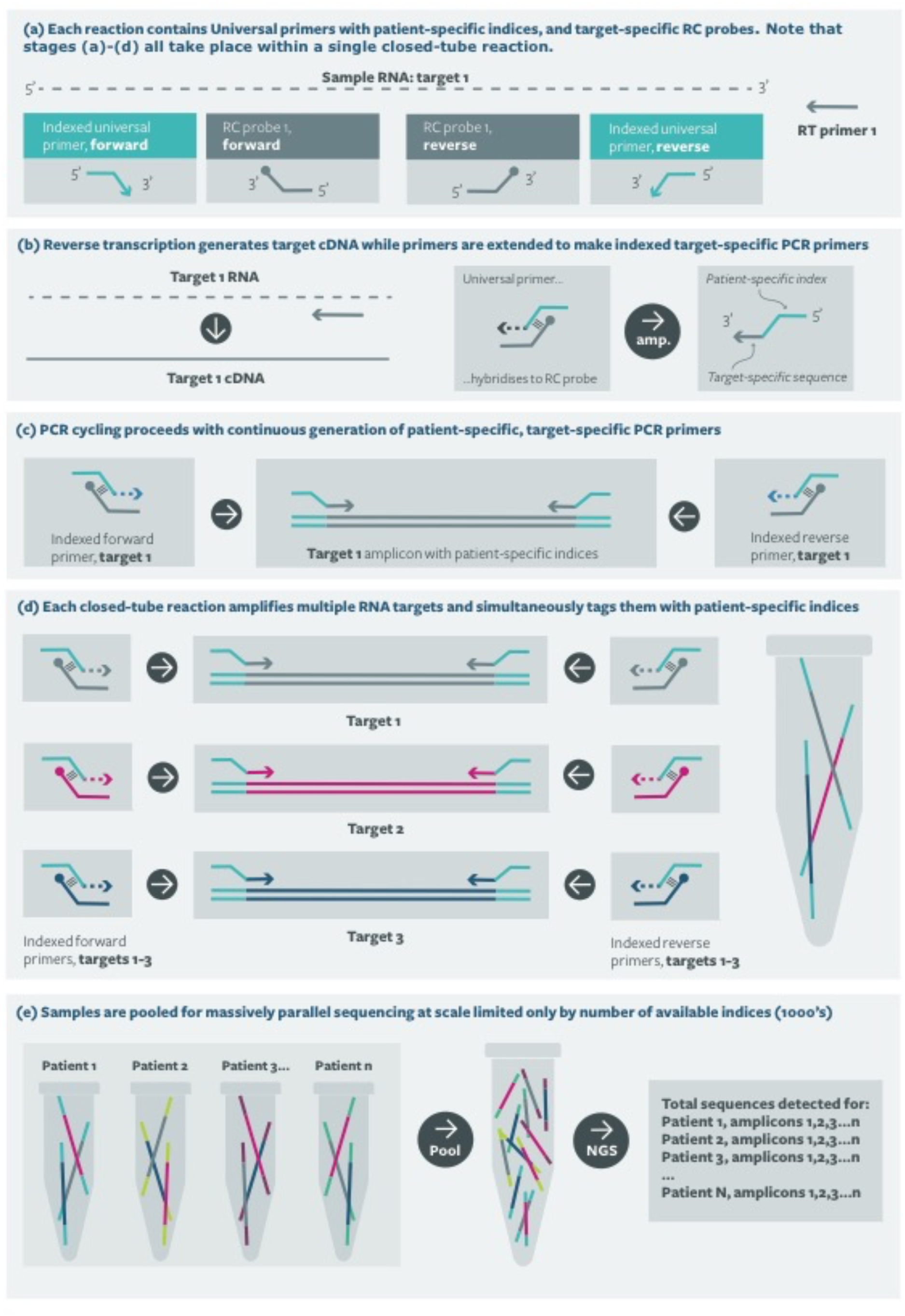
Principle of RT-RC-PCR.

The core technology is Reverse complement PCR (RC-PCR), which is a simple, robust method for uniquely indexing samples for downstream analysis in individual, closed tube reactions. The reaction is analogous to a standard PCR except that target specific primers are not provided in the reaction mix. Instead of each primer, a system comprising two oligonucleotides, a universal indexing primer (UIP) and a reverse complement probe (RC-probe) (Figure 1a), is used to generate fully formed primers as the reaction proceeds (Figure 1b). The UIP includes the necessary sequencing adaptor at the 5’ end, a sample identification sequence (10bp index sequence) in the middle and a universal sequence at the 3’ end. Note that the universal primer has no target binding sequence. The RC-probe comprises the reverse complement of the desired target binding sequence for the primer at the 5’end, and the 3’ end is able to hybridise to the universal sequence of the UIP (Figure 1b). Thus bound, the UIP can be extended, using the RC-probe as the template, to yield fully formed, indexed primers (Figure 1c), which are then available to amplify the template in subsequent rounds of the PCR. A single set of target specific probes can be used to simultaneously amplify and index many thousands of samples using an appropriate number of uniquely indexed UIPs (Figure 1d).

Combining this system with an initial reverse transcription allows direct amplification from RNA targets and sample specific indexing in a one-step, single tube reaction. For high capacity testing, the only subsequent processing required is product pooling, clean-up and Illumina sequencing (Figure 1e).

### RT-RC-PCR assay

Each RT-RC-PCR, in a final volume of 20µl, included: 1x RT-PCR mix containing reverse transcriptase and PCR polymerase; 50pM of each forward and reverse RC-probe specific to the E and RPP30 amplicons, 1nM IPs, and 5µl sample. The reaction comprised a reverse transcription step followed by 40 cycles of PCR. Amplification products were pooled and quantified by Qubit before clean-up, dilution and sequencing.

### Sequencing analysis

Whilst our assay is agnostic to sequencing platform, the critical parameter for any next generation sequencing approach is the number of reads that can be generated per given time frame. In this context Illumina sequencing provides the highest available ‘per-run’ capacity for any current platform. Our NGS analysis was performed using Illumina V2 Micro flowcells, using a dual indexing regime. Typically we analysed 200-400 samples per run; this read format and scale of analysis was purposely excessive in order to allow modelling to determine minimum read depth requirements and read length for a robust clinical test.

### Statistical analysis

The calling algorithm was developed using a training set comprising 12 independent series of Twist synthetic RNA control diluted in RNA samples that had previously tested negative by the SoC test (2000 to 0 genome copies /μl). Any sample with an aggregate of less than 100 reads assigned to the E amplicon and RPP30 amplicons combined was deemed to have failed and excluded from further analysis. All remaining analyses in the training set were subjected to simple logistic regression using the proportion of mapped reads that aligned to the E amplicon as the sole parameter (i.e. E reads / E reads + RPP30 reads, see Figure 2(d)). Logistic regression was performed using the real-statistics Excel add-in from https://www.real-statistics.com/. Baseline probability for calling a positive was set at 0.8. Diagnostic test evaluation parameters were calculated using Medcalc (https://www.medcalc.org/calc/diagnostic_test.php).

**Figure 2:**
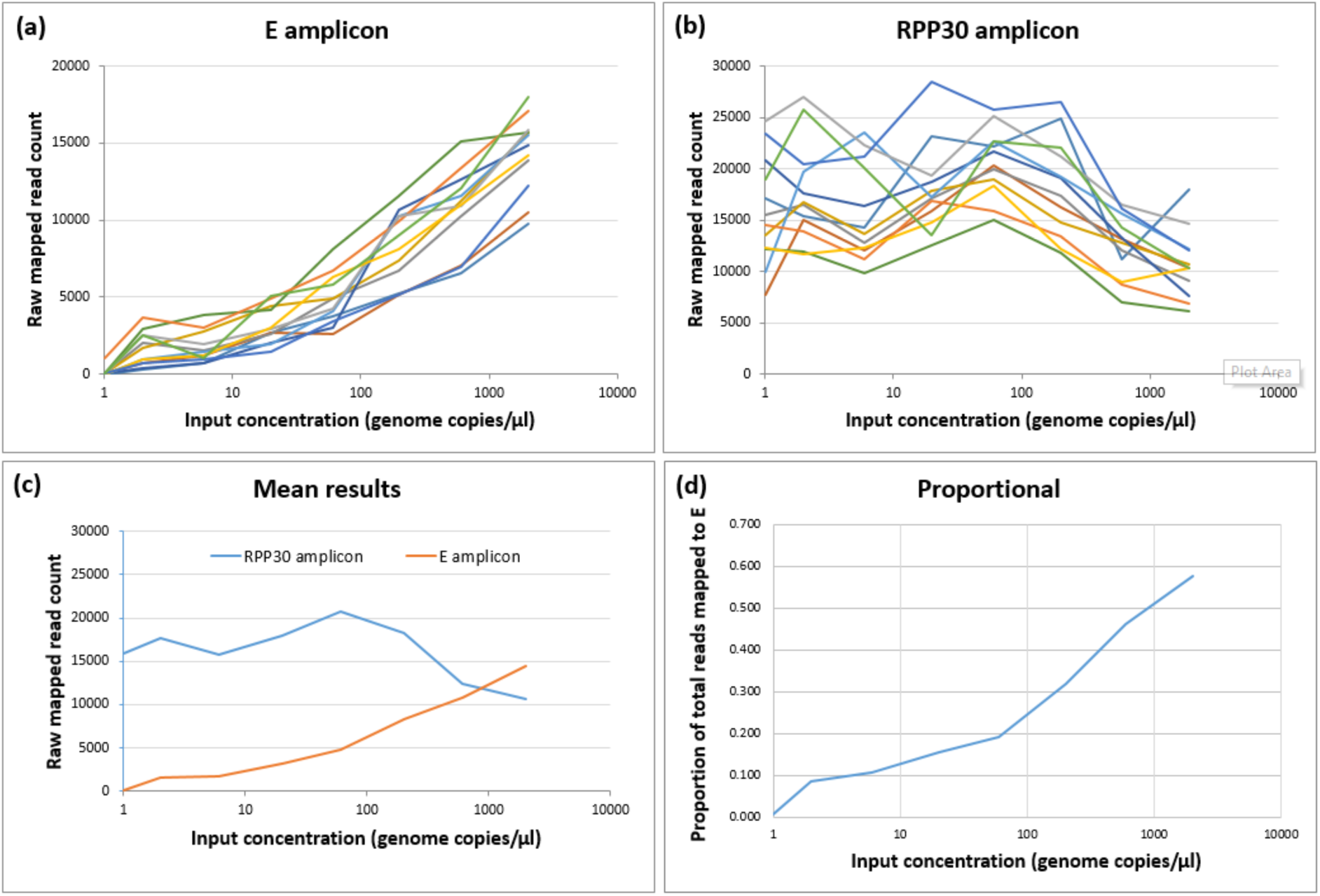
Assay performance over 12 independent serial dilutions. (a) E amplicon raw read count, (b) RPP30 amplicon (sample presence control) raw read count, (c) comparative analysis of raw read counts for both amplicons averaged across the dilutions – note the drop in RPP30 output at higher concentrations of the SAS-COV-2 RNA, (d) data shown as proportion of all aligned reads that map to the E amplicon [E / (E + RPP30)]

## Results

### Limit of detection

The limit of detection (LoD) was assessed using quantified synthetic SARS-CoV-2 RNA (Twist). This RNA was spiked into pooled negative control NPS samples and used to generate dilution series from 2000 to 0 genome copies/μl. **Figure 2** summarises the results of these analyses in terms of the raw read counts assigned to the E amplicon and the RPP30 control amplicon. As expected, the E amplicon yielded read counts proportional to the input concentration of SARS-COV-2 RNA, whilst read count for the control remained essentially consistent across the different concentrations. Some drop-off in RPP30 read count was observed where very high concentrations of SARS-COV-2 RNA were present; this can be explained by competition between the two amplicons in the diplex reaction. **Table 1** shows the qualitative detection in 12 independent dilution series using the derived calling algorithm. Dilutions showing incomplete detection across the 12 dilution series (20 copies/μl to 2 copies/μl) were used to calculate the LoD using logistic regression. Calculated LoD was 31.08 copies/μl.

**Table 1:**
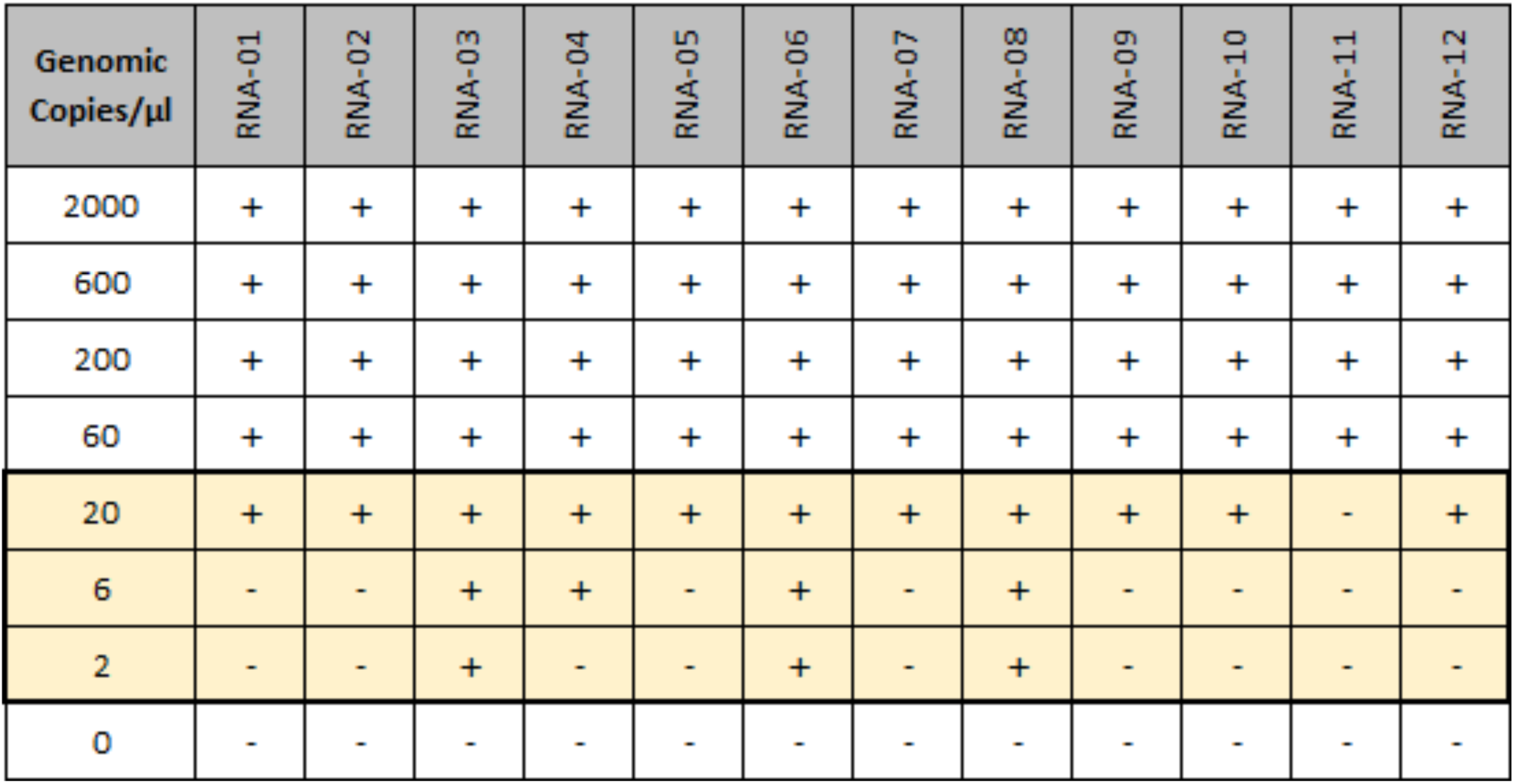
Limit of detection. Qualitative detection of artificial SARS-COV-2 RNA in 12 independent serial dilutions. Highlighted lines were used to calculate the limit of detection using logistic regression. Calculated LOD = 37.1 genome copies / μl.

| Genomic Copies/μl | RNA-01 | RNA-02 | RNA-03 | RNA-04 | RNA-05 | RNA-06 | RNA-07 | RNA-08 | RNA-09 | RNA-10 | RNA-11 | RNA-12 |
| --- | --- | --- | --- | --- | --- | --- | --- | --- | --- | --- | --- | --- |
| 2000 | + | + | + | + | + | + | + | + | + | + | + | + |
| 600 | + | + | + | + | + | + | + | + | + | + | + | + |
| 200 | + | + | + | + | + | + | + | + | + | + | + | + |
| 60 | + | + | + | + | + | + | + | + | + | + | + | + |
| 20 | + | + | + | + | + | + | + | + | + | + | - | + |
| 6 | - | - | + | + | - | + | - | + | - | - | - | - |
| 2 | - | - | + | - | - | + | - | + | - | - | - | - |
| 0 | - | - | - | - | - | - | - | - | - | - | - | - |

### Validation in extracted NP RNA

Clinical validation was carried out on RNA extracted from 192 NPS samples comprising 78 positives and 114 negatives characterised by the local rRT-PCR SoC test. **Table 2** presents the results of blinded analysis performed using RT-RC-PCR and the calling algorithm derived above. Of the 192 samples analysed, 78/78 positives called by rRT-PCR were correctly identified, two negatives were failed because the total mapped read count was less than 100 (1% overall failure rate), and 109/112 of the remaining negatives were correctly identified. Raw calculated sensitivity was 95.4-100% (95% CI) and specificity was 92.4-99.4% (95% CI). These analyses were carried out on a single Illumina V2 Micro flowcell, which represents an allocation of approximately 20,000 reads per sample. Since available reads will ultimately determine the feasible sample capacity per run, we performed random sub-sampling of these data to model performance characteristics with fewer reads available per sample. At 10 fold sub-sampling, which represents the performance with 2,000 samples on a V2 Micro flowcell run, with an average of 2,000 reads available per sample, the calculated performance metrics were virtually unchanged: Sensitivity was 95.3-100% (95% CI) and specificity was 93.5-99.8% (95% CI) with an overall failure rate of 3% (6/192).

**Table 2:** Clinical validation. (a) Raw validation metrics representing mean allocation of 20,000 reads per sample (b) Adjusted validation using 10 fold randomly subsampled data to model performance with mean allocation of 2,000 reads per sample.

**(a)**
| Test outcomes |  | rRT-PCR (Bosphore) |  |
| --- | --- | --- | --- |
|  |  | Positive | Negative |
| RT-RC-PCR | Fail | 0 | 2 |
|  | Positive | 78 | 3 |
|  | Negative | 0 | 109 |

| Statistics | Value (%) | 95% Confidence interval (%) |
| --- | --- | --- |
| Failure rate | 1.04 | 0.13 to 3.71 |
| Sensitivity | 100.00 | 95.38 to 100.00 |
| Specificity | 97.32 | 92.37 to 99.44 |
| Accuracy | 98.42 | 95.46 to 99.67 |

**(b)**
| Test outcomes |  | rRT-PCR (Bosphore) |  |
| --- | --- | --- | --- |
|  |  | Positive | Negative |
| RT-RC-PCR | Fail | 0 | 6 |
|  | Positive | 77 | 2 |
|  | Negative | 0 | 107 |

| Statistics | Value (%) | 95% Confidence interval (%) |
| --- | --- | --- |
| Failure rate | 3.13 | 1.16 to 6.68 |
| Sensitivity | 100.00 | 95.32 to 100.00 |
| Specificity | 98.17 | 93.53 to 99.78 |
| Accuracy | 98.92 | 96.17 to 99.87 |

### Validation for use with saliva samples

Significant challenges to scaled testing lie in the timely collection of samples and the work associated with RNA extraction procedures. Initially we spiked Twist synthetic SARS-CoV-2 RNA control into heat-treated negative saliva samples, compared with negative NPS samples. These experiments indicated the chemistry had good tolerance to the presence of native saliva in the reaction (**Figure 3**). To further investigate the robustness of RT-RC-PCR to native saliva, we spiked UV-inactivated SARS-CoV-2 particles (as opposed to extracted RNA) into native, untreated saliva, negative for SARS-CoV-2. The spiked samples were then left at ambient temperature (laboratory bench) for up to 48 hours (to simulate sample transit) before heat-treatment at 95°C for 5 minutes (to simulate the necessary heat-inactivation of a live sample), preparation of dilution series, and testing by RT-RC-PCR. Compared with UV-inactivated SARS-CoV-2 spiked into water, or into saliva and assayed immediately, 48 hours of ambient incubation caused no apparent reduction in detection of viral amplicon (**Figure 4a**). By contrast, 100,000 copies of synthetic viral RNA (Twist) spiked into native saliva yielded no detectable amplicon even when heat-treated and assayed immediately (data not shown).

**Figure 3:**
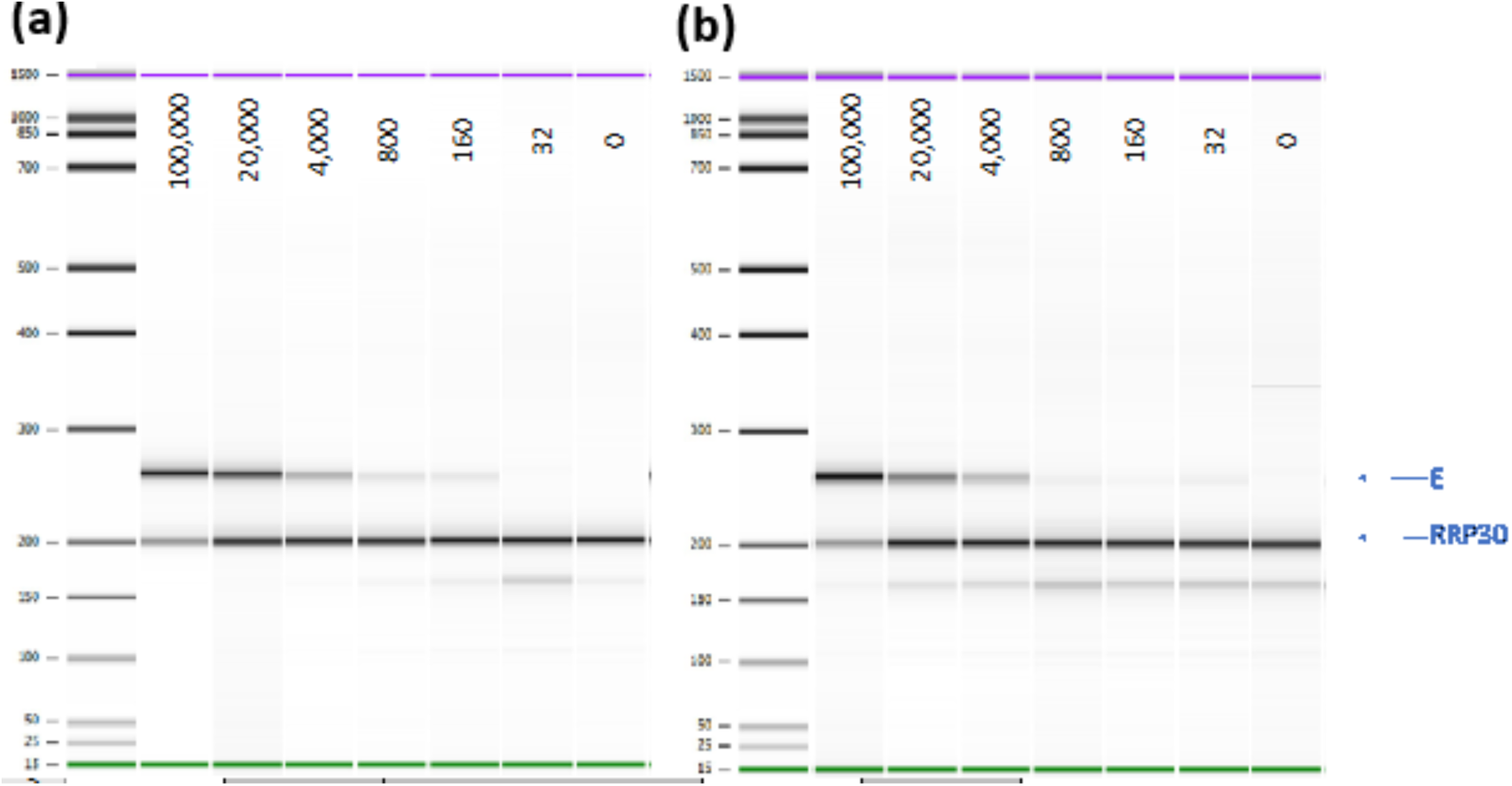
RT-RC-PCR tolerance to native saliva. The figure shows electropherograms of RT-RC-PCR amplicons derived from (a) SARS-CoV-2 synthetic RNA (Twist) spiked into extracted NPS RNA (b) SARS-CoV-2 synthetic RNA (Twist) spiked into heat-treated saliva.

**Figure 4:**
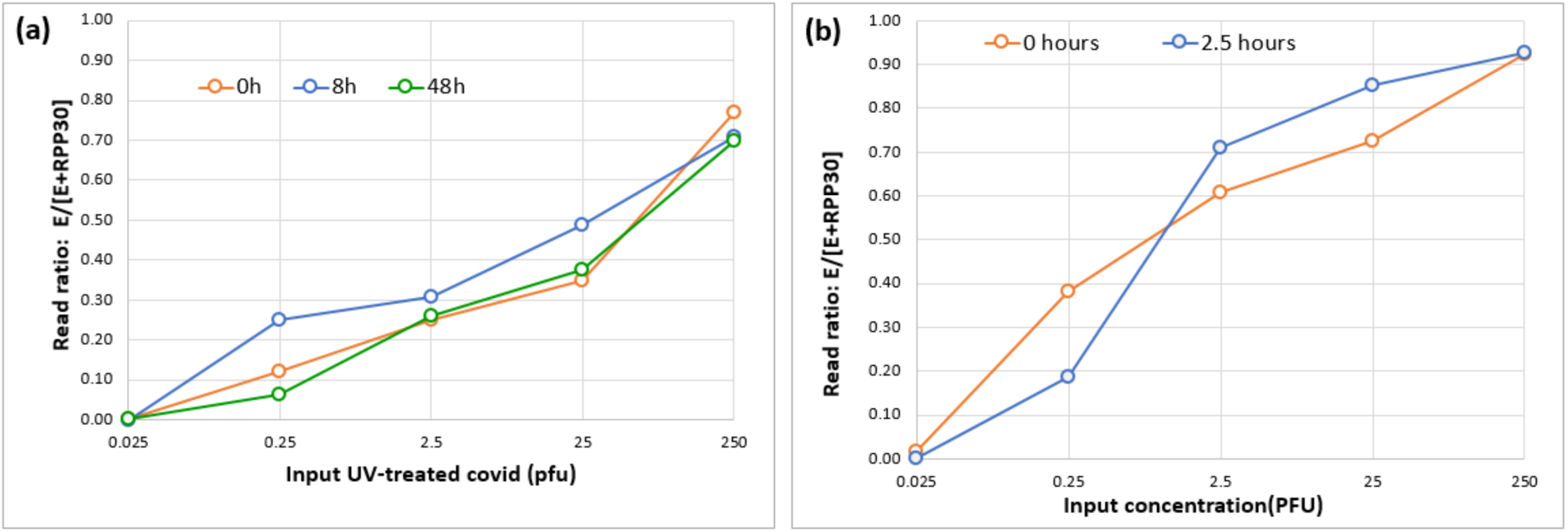
Heat treatment and delayed testing in native saliva using UV inactivated viral particles. (a) Heat treatment (95°C for 5 mins) and subsequent RT-RC-PCR at 0, 8 and 48 hours after addition of heat-inactivated virus. (b) RT-RC-PCR performed 0 hours or 2.5 hours after addition of heat-inactivated virus and heat treatment at 95°C for 5 mins.

In a final test, UV-inactivated SARS-CoV-2 was spiked into native saliva, heat-treated at 95°C for 5 minutes and then left at ambient temperature for 2.5 hours before preparation of dilution series and assay by RT-RC-PCR. Compared with a parallel dilution series assayed without delay, there was no apparent reduction in detection of viral amplicon (**Figure 4b**). Overall, these data convincingly show the robustness of RT-RC-PCR to assaying native saliva.

### Detection of variants of concern

Since our assay uses sequencing of particular parts of the SARS-CoV-2 genome as the end point measurement, we wanted to see if the assay was capable of detecting mutations and could thus have utility in typing and identifying variants of concern (VoC). Four sets of primers and RC-probes were designed to amplify regions surrounding N501Y (several VoC), delH69-V70 (UK VoC), K417N /K417T (several VoC) and E484K (several VoC).[11-14] 17 of 32 RNA samples testing positive by the local SoC rRT-PCR were shown to harbour both the N501Y and delH69-V70 mutations present in the B.1.1.7 lineage which has emerged as a UK VoC (https://assets.publishing.service.gov.uk/government/uploads/system/uploads/attachment_data/file/959438/Technical_Briefing_VOC_SH_NJL2_SH2.pdf).

## Discussion

We have developed a novel method based on RT-RC-PCR, for the qualitative detection of SARS-CoV-2 in both extracted RNA and native saliva samples. Validation studies, in comparison to rRT-PCR standard of care testing, show this method to perform robustly with high sensitivity and specificity and a low limit of detection. We have emulated a full analysis workflow using native saliva, including incubation at ambient temperature for up to 48 hours and a heat treatment viral inactivation step, with no apparent effect on performance. This testing format removes the requirement for an RNA extraction step, which is desirable in terms of cost, turn around time and availability of reagents. Interestingly, it appears the viral inactivation step performed with raw saliva essentially eliminates detection of the naked RNA control (Twist), but maintains detection of viral particles (UV inactivated control material). This suggests the test, in this form, is specific for viral particles and would not therefore detect post-infection RNA shedding, which has been noted as a potential issue in previous studies [15-18].

RT-RC-PCR has a number of major advantages in comparison to standard testing methodologies, making it suitable for laboratory based mass testing strategies.

### Safety

The single closed tube reaction reverse transcribes RNA, amplifies cDNA target material and applies unique sample identification sequences. This approach simplifies the testing process and minimises as far as possible the potential for cross-contamination. Importantly the assay also includes a multiplexed endogenous control, providing confidence in each individual assay as opposed to batch control.

### Automation

Simplicity is absolutely critical for effective high capacity automation. Our laboratory workflow is the simplest available for massively parallel analysis (PCR; Pool reaction products; Purify; Sequence). The core technology of RC-PCR is clinically validated and has been used in routine diagnostics at scale in the Wessex Genetics Service diagnostic laboratory for over 6 years.

### Sample input

RT-RC-PCR functions with extracted RNA or raw saliva without obvious loss of sensitivity. This offers an alternative to RNA extractions that often cause bottlenecks of time and availability for testing.

### Cost

List price consumable cost is approximately 2-6 GBP per sample depending on specific setup and the sequencing platform used; with economies of scale this will be highly competitive. The simplicity of the workflow means that manual intervention can be minimised, keeping staffing costs low.

### Supply chain

We validated two chemistry options from different suppliers. Neither option requires reagents currently in high demand for other methods of SARS-CoV-2 detection.

### Capacity for further development

Using a duel index system sequencing can be performed on a MiSeq instrument in 5 hours. We have performed simulations using trimmed reads and single indexing, which indicate sequencing time could be reduced to around 3 hours. We have also demonstrated that the assay can be multiplexed and can be further adapted to include other desirable targets without increasing the complexity of the lab process, or significantly affecting per-sample cost of testing. For example in the current context, amplicons could be added to allow differential diagnostics (Flu A & B, RSV). We propose that RT-RC-PCR offers future capacity for detection of other infectious diseases.

### Speed at scale

We have only analysed small numbers of samples on Illumina V2 Micro flowcells, but we have demonstrated, through subsampling our data, that accuracy is not compromised by allocating much fewer reads per sample. Conservative analysis shows that 10,000 samples could be analysed on a single Miseq V2 flowcell with no loss in sensitivity: The only constraint would be availability of suitable index sets. Other tests may have a faster, per run, measurement phase, but the realities of massively scaling tests in a laboratory make it largely unrealistic to target at-scale turnaround times, from sample to reported result, in less than 24 hours, regardless of the speed of the actual test. While RT-RC-PCR is not appropriate for point-of-care testing, it potentially adds vital capacity to population-based testing.

### Scalability

RT-RC-PCR offers the highest single run capacity available. High scaling does not require large numbers of instruments, avoiding all the difficulties of cost, space and logistics that attend methods using high numbers of ‘small’ runs. It is not necessary to ‘pool’ samples pre analysis to achieve required testing throughputs.

### Instrumentation

RT-RC-PCR uses well established, reliable and extant laboratory instrumentation, including Illumina sequencers. These technologies are available in a wide range of laboratories in both the public and private sectors.

### Detection of variants at the point of diagnosis

We have demonstrated a capacity to identify mutations in the viral genome. Currently identifying variants relies on primary detection of positive cases, followed by reflex whole genome sequencing. The process does not currently provide full coverage (at the time of writing approximately 10% of positive cases were submitted for WGS) and can take many days. With careful assay design to select appropriate VoCs and hotspots, RT-RC-PCR offers the possibility identifying known and even new mutations as part of the primary diagnostic test.

In conclusion, we have developed a novel SARS-CoV-2 test and demonstrated that its performance is broadly equivalent to rRT-PCR. The test performs robustly on native saliva samples and the workflow is extremely simple making it highly amenable to automation. Moreover, it is cheap, highly scalable and multiplexable, which offers the opportunity for more sophisticated tests. In a time when infection rates are falling and selection pressure is being exerted on the virus through application of vaccines, early detection of variants of concern will become increasingly important. We believe RT-RC-PCR has potential to facilitate accurate mass testing with concurrent detection of variants of concern.

## Intellectual property

The core chemistry of RC-PCR is IP-protected, with the patent being owned by Salisbury NHS Foundation Trust. This chemistry was developed within the NHS and has been employed for diagnostic testing by the NHS for several years. The IP is currently licensed to the Netherlands company Nimagen B. V. for research use; however, this does not impede the ability of laboratories to license this technology for diagnostic testing for SARS-CoV-2.

## Data Availability

All data discussed in the paper has been fully disclosed therein.

## Acknowledgements

The authors gratefully acknowledge the support of: Jo Harris, Microbiology Laboratory Manger SFT; James Ryan, Senior Biomedical Scientist, Microbiology, SFT; Dr Lee Phillips, Pathology Services Manger, SFT; Professor James Batchelor, University of Southampton; Dr Karl Staples, University of Southampton.

